# Translation and validation of the Cultural Competence Self-assessment Checklist of Central Vancouver Island Multicultural Society for Health Professionals

**DOI:** 10.1101/2022.03.19.22272563

**Authors:** Alexandros Argyriadis, Evanthia Asimakopoulou, Evridiki Patelarou, Ioannis Andriopoulos, Athina Patelarou, Panagiotis Paoullis, Michalis Zografakis-Sfakianakis, Agathi Argyriadi

**Author notes:** CORRESPONDENCE TO Alexandros Argyriadis, 7 Yianni Frederickou, 1036, Nicosia, Cyprus.

## Abstract

**Introduction:** The World Health Organisation emphasizes the importance of training future healthcare practitioners to practice respectful and person-centred health care. The importance of this can be demonstrated in the example of cultural competence, which has been observed to be associated with improved patient satisfaction and concordance with recommended treatment. The aim of this study was to translate and validate in the Greek language the Cultural Competence Self-assessment Checklist of the Central Vancouver Island Multicultural Society and test it in the population of health scientists in Cyprus.

**Methods:** A cross-sectional analysis took place between October 2021 and January 2022 in 300 health scientists in Cyprus using convenient sampling. The sample consisted of doctors, nurses, psychologists, social workers and physiotherapists. In order to test the questionnaire’s internal consistency reliability we used the Cronbach’s coefficient alpha.

**Results:** After the translation of the Cultural Competence Self-assessment Checklist there was a Cronbach’s alpha indication of 0.7 in all three thematic units of the checklist. 300 participants filled in the research tool, 241 women (80.3%) and 59 men (19.6). Only 2% of the sample had attended a cultural competence training before or had an expertise.

**Conclusions:** The Greek version of the Cultural Competence Self-assessment Checklist of the Central Vancouver Island Multicultural Society is a valid instrument that can be used in the Greek language referring to health scientists both in Cyprus and Greece.

## Introduction

Multiculturalism that has been developed in most of the western societies brings new necessities in the management of diversity and the inclusion of vulnerable groups in the wider social context [1, 2, 3]. Particularly in the health sector, the need to provide access to both refugees, migrants and all people belonging to the spectrum of diversity is urgent [4, 5, 6]. Intercultural education from the other side, aims at the acceptance of diversity, cultural differentiations, respect and equality [7]. Respectively, diversity studies promote the acceptance towards different perceptions, cultural values but also habits of the everyday life [8, 9]. If applied to health sciences it aims to consolidate a successful therapeutic environment, understanding in depth the needs of patients by providing holistic care with a multicultural approach [10]. According to Barett [11] Cultural Competence is the ability to participate ethically and effectively in personal and professional intercultural settings. It requires being aware of one’s own cultural values and personal desires and their implications for making respectful, reflective and evidence-based decisions, including the capacity to imagine and collaborate across cultural boundaries [12]. The lack of cultural competence in healthcare delivery has been identified by “Healthy People 2010” as a primary factor leading to health disparities [13]. The awareness that emerged from the above research studies led to the investigation of the intercultural competence of health professionals which has created a significant research trend in recent years [14, 15, 16, 17]. The correlation between cultural competence and educational level and work experience is often studied [18], as well as the importance of non-verbal communication (eye contact, grimacing gestures) and verbal (tone and volume) for successful patient-doctor/nurse communication [19]. Studies often point out the need to strengthen the institution of intercultural mediator and further training on health staff [20]. Weaknesses on health professionals were also recorded in the management of the pandemic crisis as there is an inadequacy in the provision of immigrant care and overcoming intercultural barriers [21]. A variety of scientific tools have been used to investigate the cultural competence of healthcare staff, such as the TSET (Transcultural self-efficacy tool), which consists of 83 questions and examines staff knowledge, practice and emotional background [22]. Nursing cultural competence has also been tested in a synchronous descriptive correlation study, where exposure and outcome measurement are performed at the same time, through self-administered questionnaires, based on the PES-NWI-R (revised practice environment scale of the nursing work Index) for the evaluation of the working environment. The Maslach burnout Inventory (BMI) has also been used. In other words, there is a strong intention to measure the importance and the promotion of the cultural capacity of health professionals and this research trend is constantly increasing [23]. Nevertheless, there has been limited development of scales for assessing and/or self-assessing the cultural competence of health professionals with applications in clinical settings and none so far in the Greek language.

## Materials and methods

The aim of this study was to translate and validate in the Greek language the Cultural Competence Self-assessment Checklist of the Central Vancouver Island Multicultural Society and test it in the population of health scientists in Cyprus. To this end, the research team followed the usual validation practices of research tools as described in the field of research methodology and following the steps of translation and cultural adaptation. In particular, the validity of the content, the validity of the conceptual construction and the usability of the tool were measured.

### Data Collection and Sample of the Study

The validation of the Cultural Competence Self-assessment Checklist of the Central Vancouver Island Multicultural Society took place between October 2021 and January 2022 in 300 health scientists in Cyprus using convenient sampling. The sample consisted of doctors, nurses, psychologists, social workers and physiotherapists (Table 1).

**Table 1.**
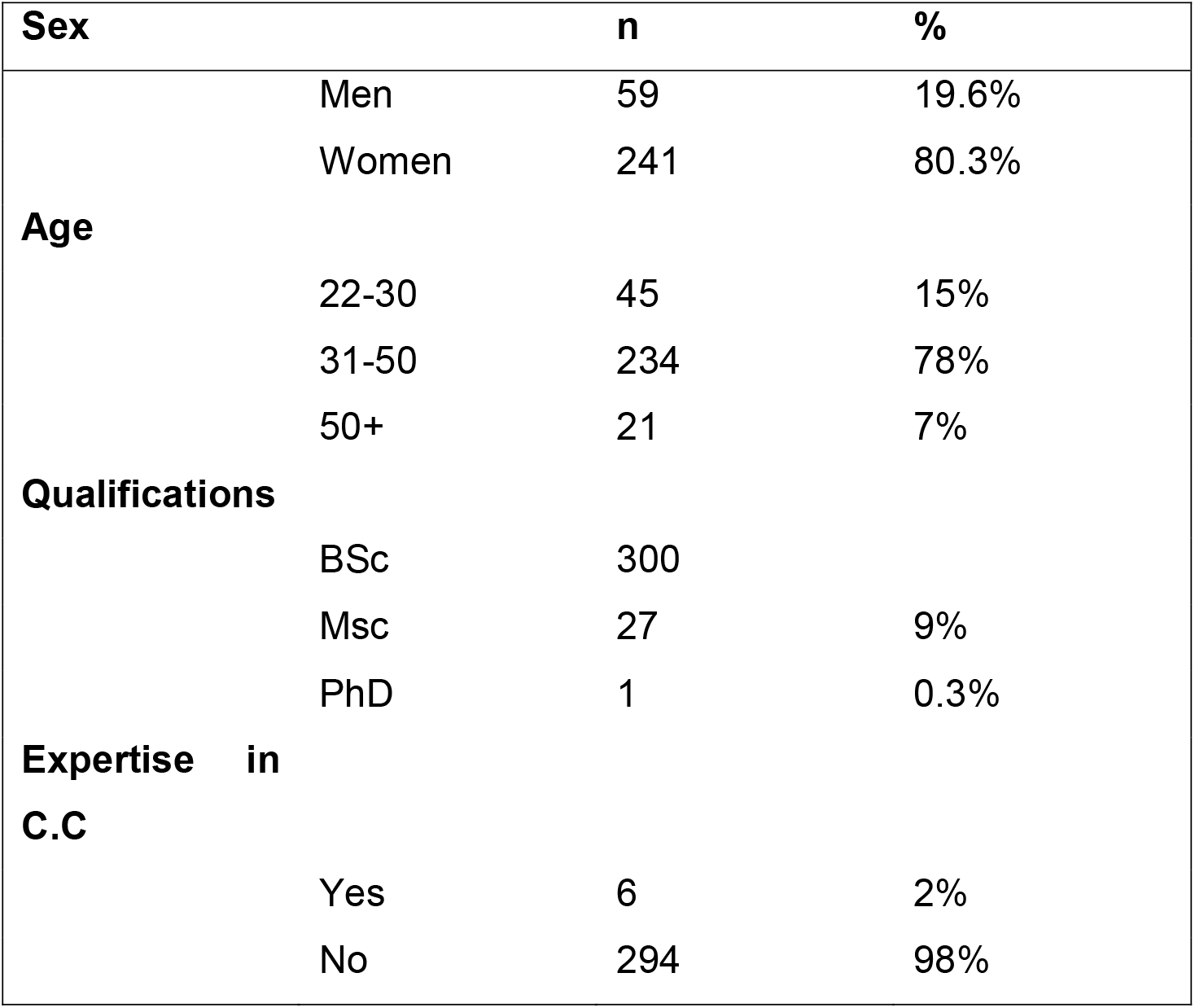
Demographic characteristics

### The questionnaire

The Cultural Competency Assessment Questionnaire was developed by the Central Vancouver Island Multicultural Society and aims to self-assess cultural competence. Its ultimate goal was to help people to consider their skills, knowledge, and awareness of themselves in their interactions with others. Its goal was to assist communities to recognize what they can do to become more effective in working and living in a diverse environment. There was a rating scale to help respondents identify areas of strength and areas that need further development in order to reach cultural competence. Although, cultural competence is a process, and learning occurs on a continuum and over a life time. This self-assessment checklist is divided into three main categories, which measure the Awareness, Knowledge and Skills. The rating of the answers is set as Never / Not at all, Sometimes / Good, Often / Fairly good and Always / Excellent with the score being respectively at the end of each section. The respondent can add up the number of times he/she has checked that column, multiple the number of times he/she has checked “Never” by 1, “Sometimes/Occasionally” by 2, “Fairly Often/Pretty well” by 3 and “Always/Very Well” by 4. The more points he/she has, the more culturally competent he/she is becoming. The first category of cultural awareness includes questions about dealing with otherness, self-knowledge, the individual’s willingness to share his or her culture and enter into a process of cultural exchange, the perception of discomfort when coming in contact with individuals from different cultural backgrounds. It also has questions about the assumptions made by individuals trying to understand the culture of another cultural group as well as questions about challenging stereotypes, reflecting on how culture influences personal judgment, behaviour and acceptance as well as possible ambiguity, curiosity and awareness of white identity. The second group that studies cultural knowledge includes questions about learning from mistakes, the assessment of knowledge, the questions that the individual asks himself in terms of cultural difference and the importance that this difference has to the individual. It also includes questions about Knowledge of History, Understanding the Impact of Culture, interest in lifelong learning and understanding the consequences of racism, sexism, homophobia, etc. Finally, this section has questions about knowledge of origin that has the individual back in time and the understanding of boundaries. The third and final section listing skills has questions about adaptability to diversity, active support for people on the diversity spectrum and intercultural communication skills. It also records the search for opportunities to acquire skills and the active involvement of the individual in processes that promote cultural experiences. Respect for diversity and the implementation of cultural practices in combination with allied strategies and flexibility are some of the necessary elements that are recorded to compose a more complete picture of the cultural ability of the individual.

### Implementation Process

The questionnaire was translated by the research team into Greek and then an inverse translation into English was followed. An experienced in health sciences bilingual translator supported the process and conducted a second independent translation to compare. Both forward-translated versions were edited and merged into the Greek version by an expert panel, using a consensus procedure. This happened in an effort to create a high quality instrument. Both in the translation phase and in the reverse translation, but also in the whole process of weighting the questionnaire, there was a continuous evaluation by an interdisciplinary seven-member team that studies the cultural competence of health professionals in Cyprus. The research team discussed the data in regularly scheduled meetings and closely monitored the process in order to evaluate possible misunderstandings and consequently to monitor the process.

Following the validation procedures, the checklist was shared in Cyprus during the period 2021-2022. The data was collected between October 2021 and January 2022 over a period of four months. This self-assessment checklist was converted into a digital questionnaire which was distributed to 375 health professionals in Cyprus electronically but the valid and complete answers amounted to 300. The process was carried out with the written consent of the participants who confirmed their desire to participate while at the same time they were informed that their personal data was secured with the anonymity provided by the specific tool. The research was approved by the Ethics Committee of Frederick University.

### Ethical Issues

For the use of this tool, the written approval of the Central Vancouver Island Multicultural Society was obtained from the scientific director of the organization. Approval was also obtained from the Frederick University Bioethics Committee as well as written consent from participants. Throughout the process anyone was able to drop out his/her participation while everyone was aware of their rights. The data collected was anonymous and ensured anonymity and respect for the personal data of the participants. All information was stored on the University premises and was used exclusively for the needs of the present study.

### Validity and Reliability

The English language version of the Cultural Competence Self-assessment Checklist of the Central Vancouver Island Multicultural Society has been tested for validity and reliability. In fact, the measurements of the Cronbach alpha values were on average 0.73, so it is already a reliable tool. During its translation into Greek, the text was checked by a philologist and the necessary corrections were made. Reverse translation followed and re-translation into Greek under the supervision of the research team of the project entitled “Building cultural competence capacity for health professionals QR-CCC” of Frederick University.

## Results

The research instrument was distributed to 375 health professionals of which 300 offered valid and complete responses. Most participants were in the age group of 31-50 years while the majority of professionals were women at a percentage of 80.3%. Only 2% of them have attended training programs in cultural studies or have some relevant specialization.

75 of the initial participants were excluded of the process because either they did not complete all the questions of the research instrument or they quit even though they had agreed to participate. The reason for quitting was their personal heavy working load and not the questionnaire, as they responded when they were personally asked. All participants were BSc holders, 27 of them (9%) had a MSc in Health Sciences and only 1 (0.3%) was a PhD holder. 294 (98%) of them had previous working experience with cultural different patients but only 6 of them (2%) had attended a relative seminar or had similar expertise. Table 1 describes analytically the demographic characteristics.

The questions of the Cultural Competence Self-assessment Checklist of the Central Vancouver Island Multicultural Society were 30 in total, divided into three thematic units of The first thematic unit that studied the cultural awareness of the participants showed high response rates of over 80% in the selection “Always / Excellent” in items A1 (91%), A2 (93.3%), and A5 (89.3%) while items A4 (66.6%), A7 (67%), A8 (68.6%) and A9 (77%) were between 60-75%. In the second thematic unit that measures knowledge in cultural issues the highest percentages over 80% were concentrated in the option “Often / Fairly Good” in the items B1 (95%) and B5 (84%). in the third thematic unit of the instrument about the skills, most answers ranged in the option “Sometimes / Good” (B9-C10) at the percentages of 60-75%. Table2 presents all results in detail.

**Table 2.** Results

| **Questions** | **Never/Not at all** |  | **Sometimes/Good** |  | **Often/Fairly Good** |  | **Always/Excellent** |  |
| --- | --- | --- | --- | --- | --- | --- | --- | --- |
|  | **n** | **%** | **n** | **%** | **n** | **%** | **n** | **%** |
| A1 | 0 | 0% | 7 | 2.3% | 20 | 6.7% | 273 | 91% |
| A2 | 0 | 0% | 3 | 1% | 17 | 5.7% | 280 | 93.3% |
| A3 | 0 | 0% | 10 | 3.3% | 255 | 85% | 35 | 11.7% |
| A4 | 1 | 0.3% | 200 | 66.6% | 76 | 25.3% | 23 | 7.7% |
| A5 | 0 | 0% | 1 | 0.3% | 31 | 10.3% | 268 | 89.3% |
| A6 | 0 | 0% | 3 | 1% | 269 | 89.7% | 28 | 9.3% |
| A7 | 0 | 0% | 198 | 66% | 89 | 29.7% | 13 | 4.3% |
| A8 | 0 | 0% | 206 | 68.6% | 77 | 25.7% | 17 | 5.7% |
| A9 | 0 | 0% | 231 | 77% | 55 | 18.3% | 14 | 4.7% |
| A10 | 83 | 27.6% | 79 | 26.3% | 100 | 33.3% | 38 | 12.7% |
| B1 | 0 | 0% | 3 | 1% | 285 | 95% | 12 | 4% |
| B2 | 0 | 0% | 6 | 2% | 230 | 76.7% | 64 | 21.3% |
| B3 | 0 | 0% | 8 | 2.7% | 17 | 5.7% | 275 | 91.7% |
| B4 | 0 | 0% | 6 | 2% | 19 | 6.3% | 275 | 91.7% |
| B5 | 0 | 0% | 6 | 2% | 252 | 84% | 42 | 14% |
| B6 | 2 | 0.7% | 10 | 3.3% | 197 | 65.7% | 91 | 30.3% |
| B7 | 8 | 2.7% | 30 | 10% | 189 | 63% | 73 | 24.3% |
| B8 | 0 | 0% | 7 | 2.3% | 203 | 67.7% | 80 | 26.7% |
| B9 | 5 | 1.7% | 195 | 65% | 30 | 10% | 20 | 6.7% |
| B10 | 9 | 3% | 200 | 66.7% | 20 | 6.7% | 71 | 23.7% |
| C1 | 4 | 1.3% | 203 | 67.7% | 46 | 15.3% | 47 | 15.7% |
| C2 | 4 | 1.3% | 209 | 69.7% | 27 | 9% | 60 | 20% |
| C3 | 4 | 1.3% | 223 | 74.3% | 31 | 10.3% | 42 | 14% |
| C4 | 7 | 2.3% | 197 | 65.7% | 74 | 24.7% | 22 | 7.3% |
| C5 | 2 | 0.7% | 199 | 66.3% | 68 | 22.7% | 31 | 10.3% |
| C6 | 2 | 0.7% | 222 | 74% | 74 | 24.7% | 2 | 0.7% |
| C7 | 1 | 0.3% | 225 | 75% | 73 | 24.3% | 1 | 0.3% |
| C8 | 3 | 1% | 231 | 77% | 62 | 20.7% | 4 | 1.3% |
| C9 | 7 | 2.3% | 195 | 65% | 67 | 22.3% | 31 | 10.3% |
| C10 | 1 | 0.3% | 204 | 68% | 82 | 27.3% | 13 | 4.3% |

### Internal Consistency

Cronbach’s Alpha indicator was used to measure the internal consistency of the instrument. As shown in table 3 the tool consists of 3 thematic units of 10 questions each. Overall, in all three thematic sections, Cronbach’s Alpha value was found to be 0.78, which demonstrates strong internal consistency. Specifically, the Cronbach’s Alpha value about the Awareness set of questions was 0.821, about Knowledge was 0.811 and about Skills was 0.712.

**Table 3.**
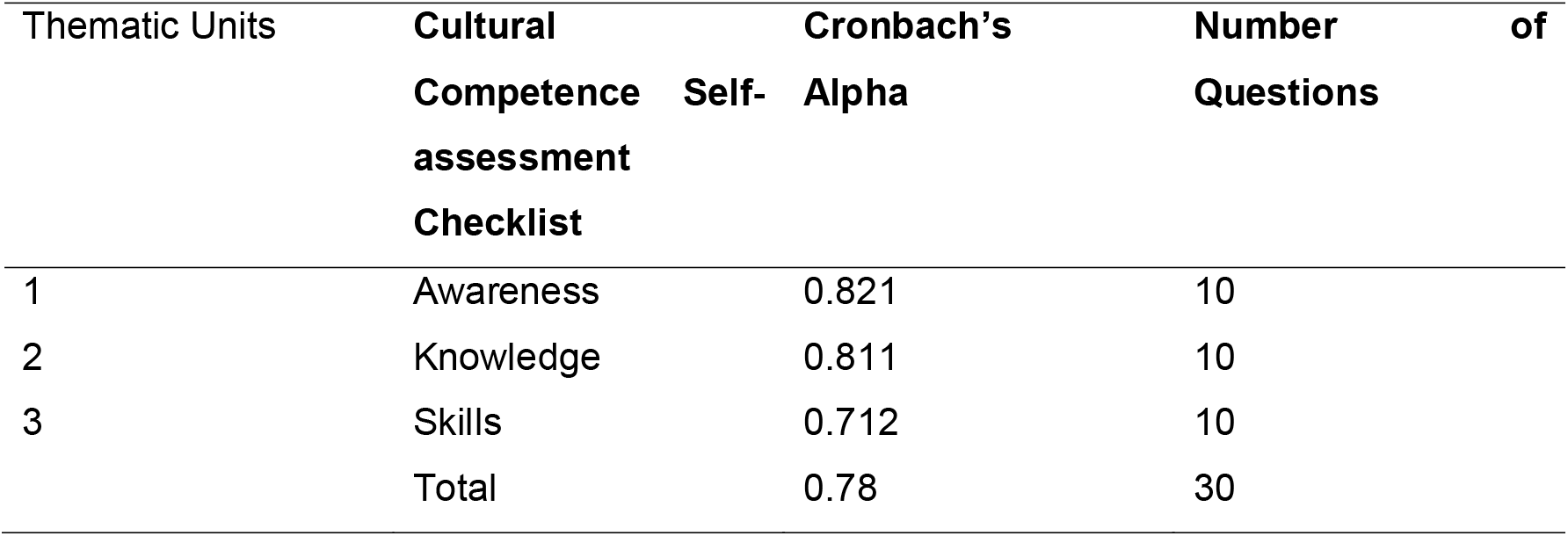
Internal Consistency Validity

### Construct Validity

In order to determine the construct validity of the instrument, we analyzed its three thematic units. The Bartletts’s results and KMO were 0.705 (KMO), x^2^= 2223, DF= 280, p < 0.001 (Bartlett) an all values were not lower than 0.5. There were not any statistically significant differences between the English and the Greek version of the instrument in any of the three thematic units. Finally, the factor analysis (F1 for Awareness, F2 for Knowledge and F3 for Skills) resulted to a low RS (0.29) and Spearman’s r = −0.046, p = 0.31 and Pearson’s r = −0.014, p = 0.798). Other variable pairs showed correlation (p < 0.05) but the absolute RS did not exceed RS which was 0.310.

### Discriminant Ability

We measured the discriminant ability of the instrument by testing the differences between the Greek and the English version. There were significant differences between sex, Awareness and Knowledge, apart from sex effect in the Skills set of questions (p = 0.177). Men showed lower mean values in Awareness and Knowledge in cultural issues related to health (17.3 ± 3.2) in comparison to women (27 ± 4.8) (p = 0.004). In contrast, the mean values in Skills and cultural competence in women were higher (29.8 ± 4.1) than in men (18.8 ± 6.2) (p < 0.001). Health professionals who had previously received training on cultural competence skills showed higher mean values (41.0 ± 5.5)

## Discussion

Taking into account the lack of research tools and self-assessment scales of cultural competence as well as the limited study in the population of health professionals, the research team for the project “Building cultural competence capacity for health professionals QR-CCC” undertook the translation and validation of the instrument in the Greek language. The English translation process and reverse translation, which was used in the present study, provided greater credibility to the linguistic validity of the Cultural Competence Self-assessment Checklist. The fact that this self-assessment checklist was converted into a digital questionnaire enhanced the ability to be answered by a sufficient number of heterogeneous sample that included participants from different cultural, educational backgrounds and experiences thus offering a better distribution. It is found that the most popular applications for publishing questionnaires on the Internet are Google Forms and Survey Monkeys [23]. Between the two applications, the Google Forms application was chosen, as there are no restrictions on designing a questionnaire using a specific application. The main advantages of using Google Forms for publishing an electronic questionnaire are the ease of creating and designing a questionnaire, without requiring programmed knowledge on the part of the researcher [24].

Measurements of internal cohesion, construct validity, discriminant ability and 3 factor-analysis were then performed, as suggested by international practices [25].

The measurements of the Cronbach alpha values of the original instrument was on average 0.73, so it was already a reliable tool. After its translation into the Greek language the Cronbach’s Alpha value was 0.78, which showed both the internal consistency of the new questionnaire and the negligible differentiation of its internal consistency from the original. In particular, in all three thematic units the internal consistency was the desired with the Cronbach’s Alpha value about the Awareness set of questions to be 0.821, about Knowledge 0.811 and about Skills 0.712. The reliability of the internal consistency of a tool’s measurements refers to the degree to which questions measuring the same feature are highly consistent or correlated, both with each other and with this feature. The reliability of this form is usually assessed through a reliability index or factor, with the Cronbach index α being the most common. Indicator values greater than 0.7 or 0.8 are usually considered satisfactory. However, it is worth noting that indicator has been severely criticized, because its implementation has strict conditions, which are difficult to meet in practice, but also difficult to assess if they are met [26].

The fact that Bartletts’s results and KMO were 0.705 (KMO), x2 = 2223, df = 280, p <0.001 (Bartlett) or all values were not lower than 0.5, showed that the construct validity of this instrument was desirable. Statistical procedures showed that no significant differences were detected between the English and Greek speaking tools.

Finally, the factor analysis performed categorizing the 3 factors into F1 for Awareness, F2 for Knowledge and F3 for Skills showed that the RS (0.29) was low and Spearman’s r was −0.046, p = 0.31 and Pearson’s r was −0.014, p = 0.798. For the successful factor analysis, the literature guidelines recommend the measurement of a variable, the creation of a correlation matrix and the choice of the method of extraction of the factors with the final step of interpretation. Exactly the same steps were followed in this study for the purpose in order to ensure an effective analysis [27].

There were significant differences between sex, awareness and knowledge, apart from sex effect in the skills set of questions (p = 0.177). Men showed lower mean values in Awareness and Knowledge in cultural issues related to health (17.3 ± 3.2) in comparison to women (27 ± 4.8) (p = 0.004). The validation of similar tools in English has shown similar results. In contrast, the mean values in Skills and cultural competence in women were higher (29.8 ± 4.1) than in men (18.8 ± 6.2) (p <0.001). Health professionals who had previously received training on cultural competence skills showed higher mean values (41.0 ± 5.5)

## Conclusions

After translation of the instrument in the Greek language, testing, statistical analysis and discussion with the scientific team we conclude that this is a reliable and useful tool that can be used by health professionals to self-assess their cultural competence. This is a high quality version that is very close to its original form and is expected to have exactly the same effectiveness. With this instrument, the level of cultural competence can be self-assessed by measuring the awareness, knowledge of the individual on cultural issues as well as the expected necessary skills that will enhance the professional effectiveness of health professionals. It can be used both by researchers as a questionnaire and by individuals themselves for the purpose of their continuous self-assessment. Through this process, the knowledge and skills of health professionals are expected to be enhanced so that they are able to address the cultural diversity both within the clinical context and in the community. In addition, they will be able to be active, to perceive the need for their continuous improvement, and to be able to distinguish both external and internal diversity.

Although this is a very reliable tool, the research team identified some limitations. Initially, it is advisable to test it on a larger number of sample in order to strengthen the belief that it is indeed characterized by high efficiency. It also needs a test and re-test over time to determine if it is valid over time. Finally, the Greek literature includes limited research data related to the self-assessment of cultural competence, while in case it is enriched with new data we will be able to strengthen the value of this tool.

## Data Availability

All data produced in the present study are available upon reasonable request to the authors

## ETHICAL APPROVAL AND INFORMED CONSENT

Ethical approval and informed consent were obtained before the implementation.

## DATA AVAILABILITY

All data are available if asked.

